# The Canadian Food Intake Screener for assessing alignment of adults’ dietary intake with the 2019 Canada’s Food Guide healthy food choices recommendations: Scoring system and construct validity

**DOI:** 10.1101/2022.12.19.22283575

**Authors:** Joy M. Hutchinson, Kevin W. Dodd, Patricia M. Guenther, Benoit Lamarche, Jess Haines, Angela Wallace, Maude Perreault, Tabitha E. Williams, Maria Laura da Costa Louzada, Mahsa Jessri, Simone Lemieux, Dana Lee Olstad, Rachel Prowse, Janis Randall Simpson, Jennifer E. Vena, Kathleen Szajbely, Sharon I. Kirkpatrick

## Abstract

The Canadian Food Intake Screener/Questionnaire court canadien sur les apports alimentaires was developed to rapidly assess alignment of adults’ dietary intake over the past month with the 2019 Canada Food Guide’s healthy food choices recommendations. From July to December 2021, adults (n=154) aged 18-65 years completed the screener and up to two 24-hour dietary recalls. The screener scoring system was aligned with the Healthy Eating Food Index-2019 (HEFI-2019), to the extent possible. ANOVA compared screener scores among subgroups with known differences in diet quality. Using the recall data, the National Cancer Institute multivariate method was used to model HEFI-2019 components, with the screener score as a covariate, and the correlation coefficient between screener and total HEFI-2019 scores was estimated. The mean screener score was 35 points (SD=4.7; maximum, 65), ranging from 25 (1^st^ percentile) to 45 (99^th^ percentile). Differences in scores in hypothesized directions were evident by gender identity (p=0.06), perceived income adequacy (p=0.07), education (p=0.02), and smoking status (p=0.003). The correlation between screener and HEFI-2019 scores was 0.53 (SE=0.12). The screener’s moderate construct validity supports its use for rapid assessment of alignment of adults’ intake with the healthy food choices recommendations when comprehensive dietary assessment is not possible.

**Novelty:**

- The Canadian Food Intake Screener was developed to rapidly assess alignment of dietary intake with Canada’s Food Guide-2019 healthy food choices recommendations.
- Scoring is aligned with the Healthy Eating Food Index-2019, to the extent possible.
- Among a sample of adults, variation in screener scores was noted, mean screener scores differed between some subgroups with known differences in diet quality, and a moderate correlation between screener scores and total Healthy Eating Food Index-2019 scores based on repeat 24-hour dietary recalls was observed.
- The Canadian Food Intake Screener has moderate construct validity for rapid assessment of overall alignment of adults’ dietary intake with the Canada’s Food Guide-2019 healthy food choices recommendations when comprehensive dietary assessment is not possible.

## Introduction

In 2019, Health Canada released an updated Canada’s Food Guide (CFG-2019), with recommendations on healthy food choices and healthy eating habits for Canadians aged two years and older (Health Canada 2022). The healthy food choices recommendations contain advice on the types of foods and beverages to eat each day and to limit (Health Canada 2022). Researchers and practitioners are often interested in assessing alignment between dietary intake and food-based dietary guidance. For example, the Healthy Eating Index-Canada enabled assessment of adherence to the 2007 Canada’s Food Guide using 24-hour recall (24HDR) or food frequency data (Garriguet 2009) for research and surveillance purposes.

Similarly, the Healthy Eating Food Index-2019 (HEFI-2019) was developed to assess alignment of intakes with the CFG-2019 healthy food choices recommendations using 24HDR data (Brassard et al. 2022a, 2022b). Sufficient construct validity of the HEFI-2019 to support its use in research and surveillance has been demonstrated (Brassard et al. 2022b).

When it is not possible to administer 24HDR or other comprehensive dietary assessment instruments, brief dietary questionnaires, informally known as screeners, can provide rapid insights into dietary intake (National Cancer Institute 2015; Thompson et al. 2015). For example, the Behavioral Risk Factor Surveillance System (Centers for Disease Control and Prevention n.d.) fruit and vegetable module has been administered in the Canadian Community Health Survey (CCHS) as an indicator of the healthfulness of dietary intake in cycles that do not involve the collection of 24HDR (Statistics Canada 2021). Recently, multi-factorial screeners have been developed to assess alignment with food-based dietary guidance in other countries (Colby et al. 2020; de Rijk et al. 2021; Gabe and Jaime 2019).

The Canadian Food Intake Screener/Questionnaire court canadien sur les apports alimentaires (Supplemental File S1) was developed to assess alignment of adult’s dietary intake over the past month with the CFG-2019 healthy food choices recommendations (Health Canada 2022). The 16-question screener is intended for self-administration by adults aged 18-65 years with marginal and higher health literacy (Hutchinson et al. 2022). In collaboration with Health Canada and external expert advisors (Supplemental File S2), guiding principles were defined to inform the screener’s development and evaluation. The final versions in English and French were informed by three rounds of cognitive testing with adults aged 18 to 65 years and face and content validity testing with a separate panel of experts, as described in the accompanying paper (Hutchinson et al. submitted). Although the CFG-2019 plate depicts the suggested proportionality of different food categories (Health Canada 2022), the screener is frequency-based in keeping with the guiding principle of a simple screener and because not all recommendations are expressed using proportionality. Given evidence of substantial gaps between the dietary intake of individuals aged two years and older and the guidance within CFG-2007 (Garriguet 2009; Hutchinson and Tarasuk 2021; Jessri et al. 2017; Tugault-Lafleur et al. 2019) and CFG-2019 (Brassard et al. 2022b), a focus on frequency of intake of healthy foods and foods to limit (Health Canada 2022) was deemed likely to differentiate higher from lower alignment with the healthy food choices recommendations.

Assessment of construct validity is important to determine whether a tool is suitable for its intended purpose, as determined by whether it performs as expected based on the theory underlying its development (Frongillo et al. 2019; Kirkpatrick et al. 2019a). The objective of this study was to assess the construct validity of the Canadian Food Intake Screener by examining variability in screener scores, whether the screener produces scores that differ between subgroups with known differences in dietary quality, and the correlation coefficient between screener scores and total HEFI-2019 scores among a sample of adults. This paper also describes the scoring system for the screener.

## Methods

Data collection was conducted online from July to December 2021. The study was reviewed by and received ethics clearance through a University of Waterloo Research Ethics Committee (ORE #42973), the University of Guelph Research Ethics Board (REB #21-04-009), and the Health Canada and Public Health Agency of Canada Research Ethics Board (REB #2020-044H). Reporting follows the *Strengthening the Reporting of Observational Studies in Epidemiology-Nutrition* (STROBE-nut) guidelines (Lachat et al. 2016).

### Reference measure and sample size calculation

In the absence of an unbiased measure of usual diet quality relative to the healthy food choices recommendations within CFG-2019, total HEFI-2019 scores, based on repeat 24HDRs, were selected as the reference measure for examining the screener’s construct validity. The HEFI-2019 has been shown to detect sufficient variation in scores reflecting alignment of dietary patterns with the healthy food choices recommendations and to differentiate between subgroups with known differences in diet quality (Brassard et al. 2022b). Additionally, HEFI-2019 scores correlate strongly with scores on the Healthy Eating Index-2015, which assesses alignment of dietary intake with the 2015-2020 Dietary Guidelines for Americans with good construct and predictive validity (Reedy et al. 2018). The evaluation of the HEFI-2019 drew upon 24HDRs (Brassard et al. 2022b), which have been shown to capture intake with less bias than other self-report dietary assessment methods, such as food frequency questionnaires (Freedman et al. 2014, 2015; Kirkpatrick et al. 2022b). The collection of repeat non-consecutive 24HDRs enables application of methods to estimate HEFI-2019 scores based on the simulated distribution of usual intakes (Zhang et al. 2011; Brassard et al. 2022b).

The sample size calculation was based on the expected correlation coefficient between screener scores and total HEFI-2019 scores, estimated using repeat non-consecutive 24HDRs. A correlation coefficient >0.4 is often considered to indicate an acceptable association, whereas coefficients of 0.5-0.7 are often observed for frequency-based dietary questionnaires (Cade et al. 2007, Willett et al. 2012) and are considered to indicate a reasonably good association (de Rijk et al. 2021; England et al. 2017; Gicevic et al. 2021; Gilsing et al. 2018; Gnagnarella et al. 2018; Schröder et al. 2011). Based on a hypothesized correlation coefficient of at least 0.4 (null hypothesis of 0.2, with an alpha of 0.05 (two-sided), 80% power), it was estimated that a sample size of 163 participants was needed. This hypothesized coefficient was based on the anticipated performance of a screener aiming to capture the multidimensional healthy food choices recommendations within CFG-2019, as well as observed correlation coefficients of other dietary screeners with various reference instruments (Colby et al. 2020; Gicevic et al. 2021; Gnagnarella et al. 2018; Schröder et al. 2011, 2012).

### Sampling and data collection

A diverse sample of adults aged 18-65 years living in Canada who could complete the screener and 24HDRs in English or French was sought. Quota sampling was used to seek one-third of the sample completing the study in French. This was a planned oversample based on the 2016 Census, which indicated that French is the first official language spoken among 21% of individuals in Canada (Government of Canada 2022). Quotas were also applied for gender identity, seeking at least 30% of participants identifying as men, given that nutrition research is often skewed toward women (Carroll et al. 2021; Colby et al. 2020; de Rijk et al. 2021; Lamarche et al. 2021); racial/ethnic identity, seeking at least 25% identifying as a racial/ethnic identity other than White; and educational attainment, seeking at least 25% who had completed college or less. Exclusion criteria included having advanced training in nutrition, following a dietary pattern related to a health condition (e.g., gluten-free, low protein, enteral or parenteral nutrition), and self-identifying as currently undergoing treatment for a condition that may alter dietary intake (e.g., disordered eating, cancer).

In the absence of a sampling frame or resources to contract a survey firm, social media was used for recruitment to avoid a convenience sample, for example, of university students. Postings on Twitter, Facebook, and Instagram described the study and invited prospective participants to visit an online questionnaire to assess their eligibility. The eligibility questionnaire was hosted on Qualtrics (Qualtrics, Provo, UT) and collected information on age, whether the potential participant resided in Canada, whether they could read and respond to questions in English or French, and characteristics associated with the exclusion criteria and quotas, noted above. Questions related to gender identity, racial/ethnic identity, and educational attainment were based on prior surveys (Hammond et al. 2022; Statistics Canada 2022). Participants who met the eligibility criteria were invited to complete informed consent and the survey if quotas were not yet filled, while those filtered out due to quotas having been achieved were thanked for their interest.

The survey, also hosted on Qualtrics (Qualtrics, Provo, UT), included the Canadian Food Intake Screener, as well as the Canadian Eating Practices Screener/Questionnaire court canadien sur les pratiques alimentaires, developed to assess adults’ alignment with the CFG-2019 healthy eating habits recommendations and described elsewhere (Haines et al., 2022).

The survey also queried sociodemographic and health characteristics, using questions informed by prior surveys (Hammond et al. 2022; Statistics Canada 2022). In appreciation of their time, participants who completed the survey received a $15 Interac e-transfer or grocery store gift card. Ongoing data quality checks were based on survey completion time (not less than 5 minutes), completing the survey only once, and correct responses to at least one of three data integrity questions, as well as peaks in survey completion and mismatches between names and email addresses that were indicators of likely bogus participants or bots (Pei et al. 2020; Storozuk et al. 2020).

Participants passing data quality checks were invited by email to complete two 24HDRs using the Canadian adaptation of the Automated Self-Administered 24-hour Dietary Assessment Tool (ASA24-2018-Canada), which can be completed in English or French (National Cancer Institute 2022a). ASA24 was developed by the National Cancer Institute (NCI) (Subar et al. 2012) and adapted to the Canadian food supply by Health Canada (National Cancer Institute 2022b). ASA24-Canada-2018 prompts participants to report everything they ate and drank from midnight to midnight on the previous day using multiple passes adapted from the interviewer-administered Automated Multiple-Pass Method (Blanton et al. 2006; Moshfegh et al. 2008), used in the nutrition-focused cycles of the CCHS (Health Canada 2006, 2017). ASA24 recall data have been shown to be comparable to those based on interviewer-administered 24HDR and to observed intake in feeding studies (Kirkpatrick et al. 2014b, 2019b). Reported foods and beverages are auto-coded by ASA24-Canada-2018 using the 2015 Canadian Nutrient File (Government of Canada 2021) and a recipe database used for surveillance by Health Canada (Health Canada 2017; National Cancer Institute 2022b). When foods in ASA24 could not be matched to existing codes in the Canadian database, US foods were used and when necessary, nutrient values were adjusted to account for fortification differences between the two countries. The assignment of food codes enables estimation of energy and nutrient values per food and per day.

The two 24HDRs were spread over approximately one month to align with the period captured by the screener, as well as across days of the week at the group level to permit accounting for potential weekend day vs. weekday effects. Although the screener captured frequency of intake during the month prior to the survey, the 24HDRs were administered after the survey to avoid the possibility that engaging in a detailed accounting of dietary intake might influence responses to the screener. As well, it was hypothesized that overall diet quality would not vary substantially over a few months (Bernstein et al. 2016). Invitations to complete the first recall were sent via email eight to ten days after completion of the survey, with up to three reminders, every two days, in cases in which the recall was not completed in response to the prior email. Invitations to complete the second 24HDR were sent eight to ten days after completion of the first 24HDR, or two to three days after sending the third reminder among those who did not complete the first recall. In appreciation of their time, participants were provided with a $10 Interac e-transfer or grocery store gift card for completion of each of the 24HDRs.

There were a total of 1484 completions of the eligibility questionnaire (**Figure 1**). Of these, 845 were determined to be bogus participants. Of the remaining 639, 485 were ineligible based on the eligibility criteria and quotas, and 154 eligible participants completed the survey. A total of 95 participants completed the survey in English and 59 in French. Cleaning procedures and sample sizes for the 24HDRs are detailed below.

**Figure 1:**
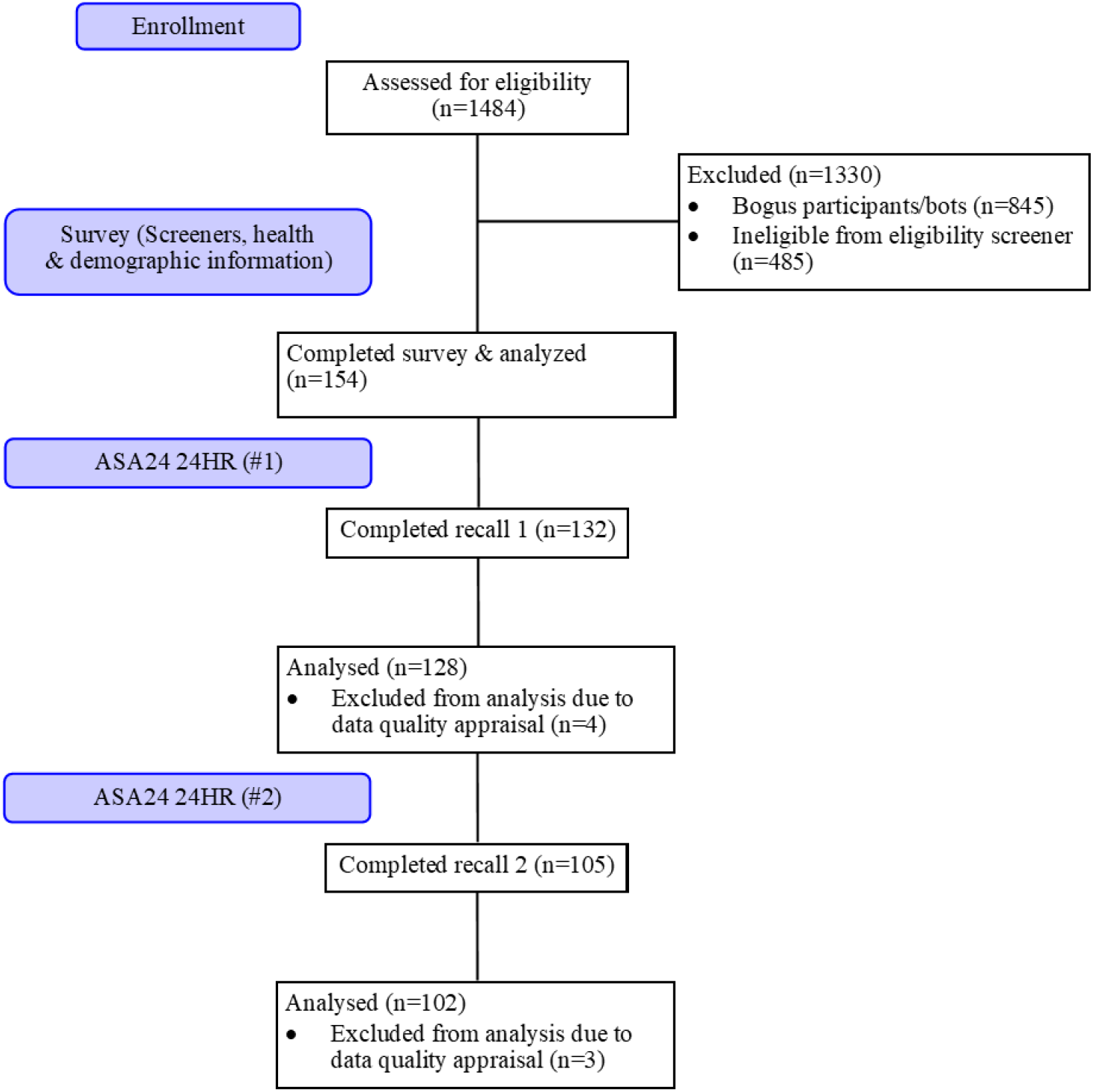
Flowchart describing participation in a study to assess the construct validity of the Canadian Food Intake Screener after applying inclusion and exclusion criteria.

### Canadian Food Intake Screener scoring system

The screener assesses frequency of intake over the past month (i.e., “Over the past month, how often did you eat…”) of healthy foods and foods to limit, according to the recommendations (Health Canada 2022). Respondents choose from 10 response options, adapted from the Dietary Screener Questionnaire and the Diet History Questionnaire (National Cancer Institute 2021; National Cancer Institute 2022c; Millen et al. 2006; Subar et al. 2001; Thompson et al. 2017), ranging from never to six or more times per day (Hutchinson et al. 2022). As a first step in scoring, the response options for each question were scored ordinally, using a scale of zero to nine (Gicevic et al. 2021; Wakimoto et al. 2006). In keeping with the scoring approaches for the HEFI-2019 (Brassard et al. 2022a, 2022b) and HEI-2015 (Krebs-Smith et al. 2018), responses to questions that capture healthy foods were scored positively (i.e., more frequent consumption was assigned a higher score), while responses to questions that capture foods to limit were reverse scored (i.e., less frequent consumption was assigned a higher score).

Next, prior to data analyses, a scoring system was developed (**Table 1**), informed by discussions with Health Canada and the advisors. The system was aligned as closely as possible with the HEFI-2019 scoring algorithm (Brassard et al. 2022a, 2022b) by using scores assigned to individual questions in combination in some cases (i.e., through summing and ratios), as well as through weighting. In the development of the screener, an effort was made to avoid combining different types of foods (e.g., meats, cheese, and milk; foods and beverages) in a single question to minimize cognitive load (Hutchinson et al. 2022). Further, based on cognitive testing, the screener includes a question specific to potatoes to help respondents report frequency of intake of both vegetables and grain foods more accurately (Hutchinson et al. 2022). Accordingly, in four cases, scores assigned to separate questions were summed to create “components”, reflecting intake of total vegetables (including potatoes) and fruits, total protein foods, foods and beverages high in sugar, and foods and beverages high in sodium/saturated fat (Table 1). The inclusion of separate questions for animal- and plant-based proteins and for whole grains and refined grains enabled derivation of ratios to reflect emphases within the CFG-2019 healthy food choices recommendations (Health Canada 2022).

**Table 1:** Scoring system for the Canadian Food Intake Screener.

| Component<sup>1</sup> | Screener questions | Scoring approach | Maximum points | Standard for minimum score (zero) | Standard for maximum score |
| --- | --- | --- | --- | --- | --- |
| Vegetables and fruits | Vegetables + potato + fruits | Responses scored 0-9 points<sup>4</sup> and summed, /27 | 20 | Never consuming vegetables, potatoes, or fruit | Consuming each item 6 or more times per day |
| Whole-grain foods | Whole-grain foods | Response scored 0-9 points | 5 | Never consuming whole-grain foods | Consuming whole-grain foods 6 or more times per day |
| Grain foods ratio | Whole-grain foods/Total grain foods<sup>2</sup> | Ratio of frequencies | 5 | Never consuming whole-grain foods | Ratio of 1.0 |
| Protein foods | Plant-based protein foods + yogurt, cheese, kefir + animal-based protein foods + unsweetened milk | Responses scored 0-9 points and summed, /36 | 5 | Never consuming protein foods | Consuming each item 6 or more times per day |
| Plant-based protein foods | Plant-based protein foods/Total protein foods<sup>3</sup> | Ratio of frequencies | 5 | Never consuming plant-based protein foods | Ratio of 1.0 |
| Unsaturated oils | Margarine and vegetable oils | Response scored 0-9 points | 5 | Never consuming margarine and vegetable oils | Consuming margarine and vegetable oils 6 or more times per day |
| Foods and beverages high in sugars | Sweetened milk + sweetened beverages + sugary snacks | Responses reverse-scored 0-9 points and summed, /27 | 10 | Consuming each item 6 or more times per day | Never consuming sweetened milk, sweetened beverages, or sugary snacks |
| Foods high in sodium/saturated fat | Fast food + processed meat + salty snacks | Responses reverse-scored 0-9 points and summed, /27 | 10 | Consuming each item 6 or more times per day | Never consuming fast food, processed meat, or salty snacks |

<sup>1</sup>Components are derived only for calculating total screener scores. Individual component scores should not be used on their own as the Canadian Food Intake Screener is intended to provide rapid insights into *overall* alignment with the Canada's Food Guide-2019 healthy food choices recommendations.
<sup>2</sup>Total grain foods = whole-grain foods + refined grain foods
<sup>3</sup>Total protein foods = animal-based protein foods + plant-based protein foods + yogurt, cheese, kefir + unsweetened milk
<sup>4</sup>Respondents to the screener choose from 10 response options for each question, ranging from never consuming to consuming the given foods or beverages six or more times per day. As a first step in scoring, the response options for each question are scored continuously, using a scale of zero to nine points, with reverse-scoring for responses to questions that capture foods to limit (i.e., less frequent consumption is assigned a higher score).

Specifically, ratios of scores assigned to frequencies were used for protein foods (plant-based protein foods/total protein foods) and grain foods (whole-grain foods/total grain foods). Scores based on frequency of intake of whole-grain foods and unsaturated oils round out the scoring system, for a total of eight components.

Weighting of the components was informed by the HEFI-2019 scoring algorithm (Brassard et al. 2022a, 2022b), with vegetables and fruits receiving 20 possible points; whole grain foods, 5 possible points; grain foods ratio, 5 possible points; protein foods, 5 possible points; plant-based protein foods, 5 possible points; unsaturated oils, 5 possible points; foods and beverages high in sugars, 10 possible points; and foods and beverages high in sodium/saturated fat receiving 10 possible points (Table 1). The weighting for vegetables and fruits, grain foods, and protein foods is consistent with the CFG-2019 plate proportions of 50%, 25%, and 25%, respectively (Health Canada 2022). Further, weighting of components capturing foods and beverages to limit was aligned (20/65 possible points, 31%) with the HEFI-2019 scoring (25/80 possible points, 31%) (Brassard et al. 2022a, 2022b). Consistent with the guidance (Health Canada 2022), no upper limit on frequency was applied in determining the standards for maximum scores for healthy foods (Table 1). The standard for minimum scores for healthy foods is zero frequency of consumption. For sources of added sugars, saturated fat, and sodium, the standard for full points is zero frequency of consumption, whereas frequencies between the minimum and maximum frequencies (Table 1) receive fewer points, consistent with the recommendations to limit these foods and beverages (Health Canada 2022).

Although the scoring process results in scores for screener “components”, these are used solely to calculate total scores. Given its brevity, the screener is designed to provide a single score indicative of higher or lower alignment with the CFG-2019 healthy food choices recommendations overall. The total possible score is 65. Code for applying the scoring system is available in the supplementary material (**Supplemental Files S3 & S4**).

### ASA24-Canada-2018 data and variables to calculate HEFI-2019 scores

Of the 154 participants in the analytic sample, 132 completed the first 24HDR and 105 completed the second. The median number of days between the initial survey and the first recall was nine (interquartile range, 8-11 days) and between the two recalls was nine (interquartile range, 8-10 days). The coded dietary intake data from ASA24-Canada-2018 were reviewed by a registered dietitian (JMH), following protocols outlined by NCI (National Cancer Institute 2022d) and with input from a second registered dietitian (SIK) and a Health Canada nutritionist experienced in coding and cleaning 24HDR data. Based on this review, four first 24HDRs and three second 24HDRs were removed because they were completed in less than five minutes (n=2), were incomplete (n=1), or had low or high estimated energy intakes (≤600 kcal or ≥4400 kcal for women and ≤650 kcal or ≥5700 kcal for men) combined with an indication by the participant that intake on the recalled day was usual (n=4). This resulted in 128 first recalls (83% of the analytic sample) and 102 second recalls (66%) included in the analyses.

Known ASA24 database issues identified by NCI (National Cancer Institute 2022e) were manually corrected, and portion size outliers were investigated and adjusted in two instances.

For example, a respondent reported consuming 6.25 cups of potato salad. This was decreased to three cups, reflecting a more plausible portion size but also that the participant selected greater than the largest option, more than two cups. The default food codes applied by ASA24-Canada-2018 to free-text entries, entered when a participant indicated they could not find the food they were looking for, were investigated and corrected in cases in which they did not match (Zimmerman et al. 2015). For example, a code for chicken breast was assigned to a free-text entry of burrito bowl, based on the food category indicated by the respondent, and manually adjusted to burrito. Corrections to free-text entries caused changes to 116 food codes (1.8% of all food codes in the dataset).

The cleaned intake data were used to estimate total HEFI-2019 scores based on the simulated distribution of usual intake, as described below. The HEFI-2019 comprises 10 components, including Vegetables and fruits, Whole-grain foods, Grain foods ratio, Protein foods, Plant-based protein foods, Beverages, Fatty acids ratio, Saturated fats, Free sugars, and Sodium (Brassard et al. 2022a, 2022b), each scored using a ratio. To arrive at scores, foods and beverages reported must be classified and amounts in appropriate units determined for the numerator and denominator for each component.

For five of the ten HEFI-2019 components, the numerators and denominators are based on reference amounts, which represent the amount of food typically consumed at one time (Health Canada 2016). A database developed by Health Canada specifying the reference amounts for food codes applied to the 2015 CCHS-Nutrition data, which was used for evaluation of the HEFI-2019 (Brassard et al. 2022b), was drawn upon. Unmatched food codes were identified, and reference amounts were assigned by Health Canada staff. Reference amounts were then merged into the ASA24-Canada-2018 data by food code, the amounts reported were divided by the reference amount for each dietary constituent of interest, and the resulting quantities were summed to arrive at total reference amounts consumed of the categories of interest, such as Vegetables and fruits, per recall. In some cases, it was necessary to disaggregate recipes into ingredients, assign reference amounts at the level of ingredients based on their proportions within the recipe, and recompile the recipe and associated reference amounts. Recipe information underlying the ASA24-Canada-2018 system was used for this purpose. The denominator for Vegetables and fruits, Whole-grain foods, and Protein foods is total foods expressed as reference amounts, computed by summing the HEFI-2019 Vegetables and fruit component, Whole-grain foods component, non-whole grain foods (used in the Grain foods ratio), animal protein foods (used in the Protein foods component), Plant-based protein foods component, unsweetened milk and unsweetened plant-based protein foods (used in the Beverages component), plus an “other” foods component that captures foods not otherwise included in HEFI-2019 components, such as condiments, processed meats, and desserts. Total foods does not include culinary ingredients, beverages without protein, and oils and spreads (Brassard et al., 2022a).

The Beverages component of the HEFI-2019 is computed as the ratio of grams of water (including carbonated water), unsweetened milk, and unsweetened plant-based beverages containing protein to grams of all beverages, including alcohol. Except for the numerator for Free sugars, the remaining numerators and denominators–unsaturated fats, saturated fats, sodium, and energy–were available from the ASA24-Canada-2018 data. The process to calculate amounts of free sugars per recall was similar to that for reference amounts, with information on the free sugars content (per 100g) for each food code obtained from a Health Canada database (Rana et al. 2021) and estimates for codes for which free sugar values had not been determined provided by Health Canada staff.

### Sociodemographic and health characteristics

Sociodemographic and health characteristics of interest were based on known differences in diet quality observed in other studies (**Table 2**) (Alkerwi et al. 2017; Brassard et al. 2022b; Guenther et al. 2014; Hiza et al. 2013; Kuczmarski et al. 2016; Moubarac et al. 2017; Reedy et al. 2018; Tarasuk et al. 2010). Gender identity categories included men, women, and non-binary. Age was captured as a five-level categorical variable. Perceived income adequacy was assessed by asking participants, “Thinking about your total monthly income, how difficult is it for you to make ends meet?”, with response options including very difficult, difficult, neither easy nor difficult, easy, and very easy (Litwin and Sapir, 2009). For examination of known differences, response categories were condensed to a three-level variable, including difficult or very difficult, neither easy nor difficult, and easy or very easy, based on the observed frequencies. Educational attainment was captured as high school graduate or less; some college; college graduate/Collège d’enseignement général et professionnel (CÉGEP), apprenticeship training, and/or trade programs; some university; university graduate; or postgraduate training or professional degree. For examination of known differences, these categories were collapsed to two, college graduate/CÉGEP/apprenticeship training/trade program or less versus higher educational attainment. Health literacy was assessed using the Canadian adaptation of the Newest Vital Sign©, which asks six questions referencing a nutrition label (Mansfield et al. 2018). The original tool demonstrated reasonable criterion validity and reliability (Weiss et al. 2005), and the Canadian adaptation demonstrated face validity with Health Canada experts (Mansfield et al. 2018). Each question was scored as correct (1) or incorrect (0), and scores were summed to a possible six points (Weiss et al. 2005). For examination of known differences, a binary variable was created to indicate low or limited health literacy (score of 0-3) or adequate health literacy (score of 4-6) (Weiss et al. 2005). Finally, smoking status was assessed by asking participants whether they currently smoked tobacco (Statistics Canada 2021), and a binary variable was created to indicate those who currently smoked versus those who did not for examination of known differences.

**Table 2:** Hypothesized differences in Canadian Food Intake Screener scores across subgroups based on known differences in diet quality.

| **Characteristic** | **Hypothesized association** | **References** |
| --- | --- | --- |
| **Age** | Higher screener scores among older versus younger adults. | Brassard et al. 2022b; Guenther et al. 2014; Hiza et al. 2013; Moubarac et al. 2017; Reedy et al. 2018 |
| **Gender** | Higher screener scores among women versus men. | Brassard et al. 2022b; Guenther et al. 2014; Hiza et al. 2013; Moubarac et al. 2017; Reedy et al. 2018 |
| **Income adequacy** | Higher screener scores among individuals with higher versus lower income adequacy. | Tarasuk et al. 2010; Hiza et al., 2013 |
| **Educational attainment** | Higher screener scores among individuals with higher versus lower educational attainment. | Tarasuk et al. 2010; Hiza et al. 2013 |
| **Health literacy** | Higher screener scores among individuals with higher versus lower health literacy. | Kuczmarski et al. 2016 |
| **Smoking status** | Higher screener scores among individuals who report not smoking versus those who report smoking. | Alkerwi et al. 2017; Brassard et al. 2022b; Guenther et al. 2014; Reedy et al. 2018 |

### Statistical analyses

Statistical analyses were conducted using SAS, version 9.4 (SAS Institute, Cary, NC, 2013). Descriptive statistics, including frequencies, were generated to describe the sample. The assessment of construct validity included examining the variability in screener scores, comparing screener scores among subgroups with known differences in diet quality, and estimating the correlation coefficient between screener scores and total HEFI-2019 scores.

The mean screener score and standard deviation were computed. ANOVA was used to compare screener scores between subgroups defined by characteristics known to differentiate dietary quality. Levene’s test was used to assess whether the assumption of homogeneity of variances was not met, and Welch’s ANOVA was used when it was not. Individuals with non-binary gender identities and those with missing responses to the gender identity question were not included in the assessment of known differences by gender identity due to small cell sizes (<5).

The approach used in the evaluation of the HEFI-2019 (Brassard et al. 2022b) was extended to incorporate the screener score as a covariate, enabling examination of relationships between screener scores and total HEFI-2019 scores. The multivariate Markov Chain Monte Carlo (MCMC) extension (Zhang et al. 2011) to the NCI method for estimating distributions of usual intake was used to account for within-person random error in the 24HDR data. The NCI method incorporates transformations and back-transformations to address skewness and non-normality that commonly affect dietary intake data (Tooze et al. 2006, 2010). The MCMC extension allows joint modelling of the numerators and denominators used to arrive at HEFI-2019 component and total scores, including dietary components consumed nearly daily by most persons and those consumed episodically (Zhang et al. 2011). Whole and non-whole-grain foods, animal- and plant-based protein foods, beverages, and “other” foods were modelled as episodically consumed based on ≥5% of recalls not including the given category (Krebs-Smith et al. 2010). All other dietary components were considered to be non-episodically consumed. In addition to the screener score, covariates included gender identity and age, as well as 24HDR sequence (first versus second recall) and whether the 24HDR was completed for a weekday versus weekend day (Nusser et al. 1996). Inclusion of the latter two covariates allows statistical adjustment for these “nuisance effects” (Kirkpatrick et al. 2022a). These analyses made use of data for all participants with 24HDRs (Brassard et al. 2022b).

The NCI method generates simulated true usual intakes, which allow estimation of the distribution of usual intakes, conditional on individual-level covariates while maintaining the joint distribution of these covariates as observed in the sample. The generated output included scores based on the application of the HEFI-2019 scoring algorithm to simulated usual intakes, along with the screener scores used in the modeling of each HEFI-2019 component. This output, therefore, captured the relationships between the screener score and each HEFI-2019 component score, as well as the sum of the component scores, the total HEFI-2019 score.

Using this output, the Pearson correlation coefficient between screener scores and total HEFI-2019 scores was computed as the square root of the R-squared of a linear regression model, with screener scores as the independent variable and total HEFI-2019 scores as the dependent variable. This assessment of the correlation coefficient was the primary focus of the study and used to justify the sample size. The square of the correlation coefficient, or the coefficient of determination (Kasuya 2018), is the proportion of variance in the dependent variable explained by the linear relationship with the independent variable. Regression diagnostics based on the model residuals did not suggest gross violations of the linear model assumptions. Standard errors for statistics based on the NCI method were obtained using the bootstrap procedure (Tooze et al. 2010, Bland and Altman, 2015), with 200 replicates.

To assess whether higher screener scores were driven by higher overall intake, the correlation between screener scores and simulated usual energy intake based on the 24HDRs was assessed using the output from the multivariate modelling of the numerators and denominators (including energy) used to arrive at HEFI-2019 scores, with the screener score as a covariate. The Pearson correlation coefficient between screener scores and simulated usual energy intake was computed as the square root of the R-squared of a linear regression model, with screener scores as the independent variable and energy intake as the dependent variable.

For comparison, the R-squared is provided for the linear regression model that included the scores for the components generated in the process of scoring the screener, rather than the total screener score. Additionally, the implications of relaxing the standards for maximum scores for healthy foods and minimum scores for foods to limit were considered by comparing the correlation coefficients and R-squared estimates between models based on total scores derived from the original scoring system, described above, and based on an altered system that used truncation in the process of scoring. For example, for assigning minimum scores for foods to limit, such as processed meats, the response options were reverse scored continuously from zero to seven rather than zero to nine, with a score of seven applied to the three highest frequency categories. When truncation was applied to the plant-based protein component, scored as a ratio to total protein, the standard for the maximum score was 0.5, consistent with the HEFI-2019 thresholds (Brassard et al. 2022a, 2022b); whereas without truncation, the standard for the maximum score was 1.0. Further, to assess whether the *a priori* scoring system added value in aligning screener scores with total HEFI-2019 scores, and thus, with the recommendations, the estimated correlation coefficient and R-squared were compared to those estimated using screener scores based on totaling the scores from zero to nine points applied to each question, without creating components by summing across screener questions or deriving ratios.

Finally, to allow comparison with the results of the HEFI-2019 evaluation (Brassard et al. 2022b) the NCI method was rerun without the screener score included to estimate HEFI-2019 scores in the sample based on simulated usual intakes.

P-values and standard deviations/standard errors are reported along with an interpretation of the level of evidence (e.g., moderate, strong) (Muff et al. 2022).

## Results

Of the 154 participants who completed the screener, 74% identified as women and one quarter identified as men (**Table 3**). Participants were fairly evenly distributed by age categories, with the smallest proportion (10%) in the 55-65-year-old age category. Over half of participants (60%) identified as White, 13% as South Asian, 11% as East/Southeast Asian, and 6% as Black. Most participants stated it was neither easy or difficult (31%), easy (26%), or very easy (23%) to make ends meet. Most participants had completed some post-secondary education, with 16% graduating from college/CÉGEP or an apprenticeship training program or trade program, 14% and 34% completing some or graduating from university, respectively, and 24% completing postgraduate training or a professional degree. The vast majority (91%) reported they did not currently smoke. Characteristics of participants who completed 24HDRs versus those who did not were similar (data not shown).

**Table 3:**
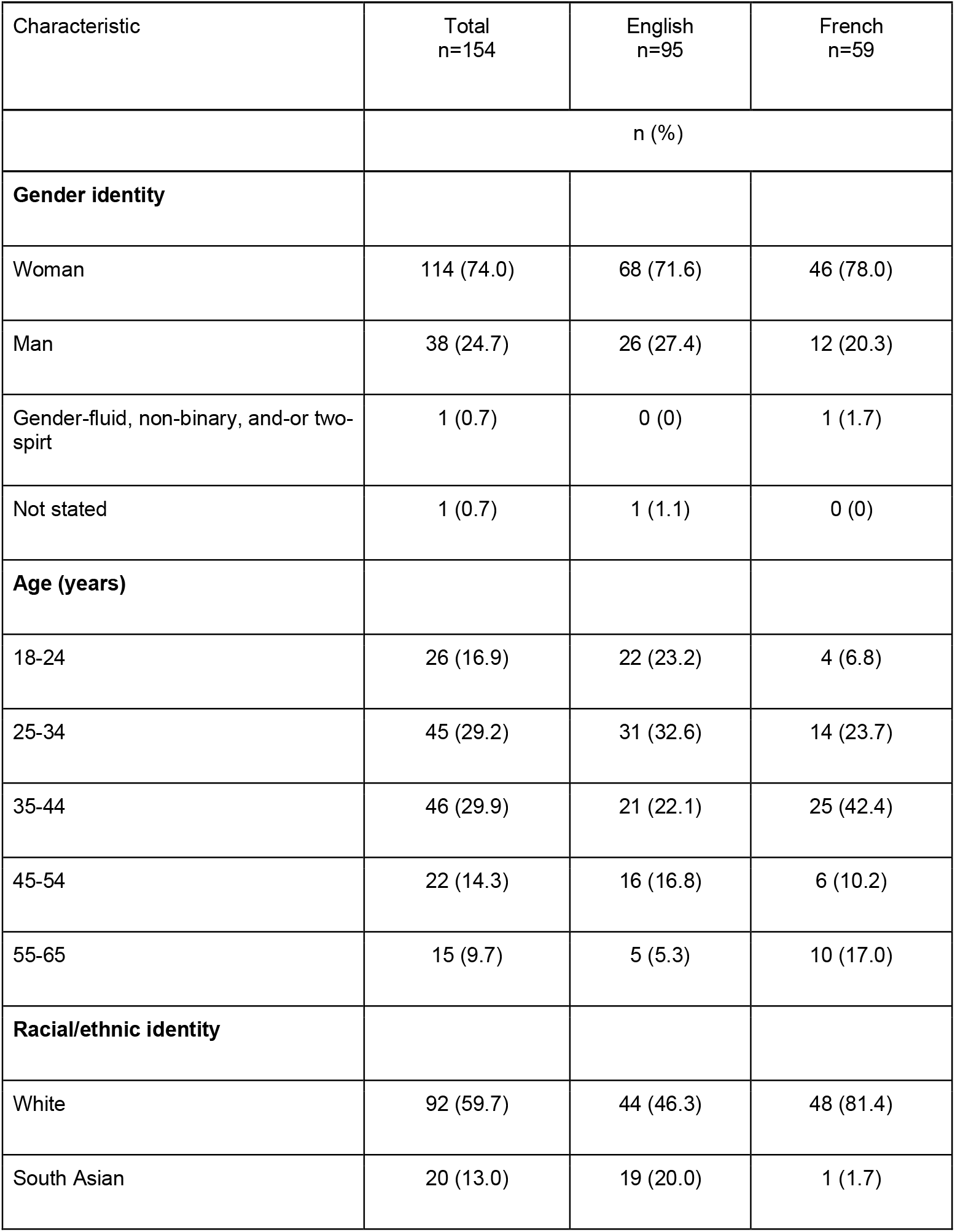
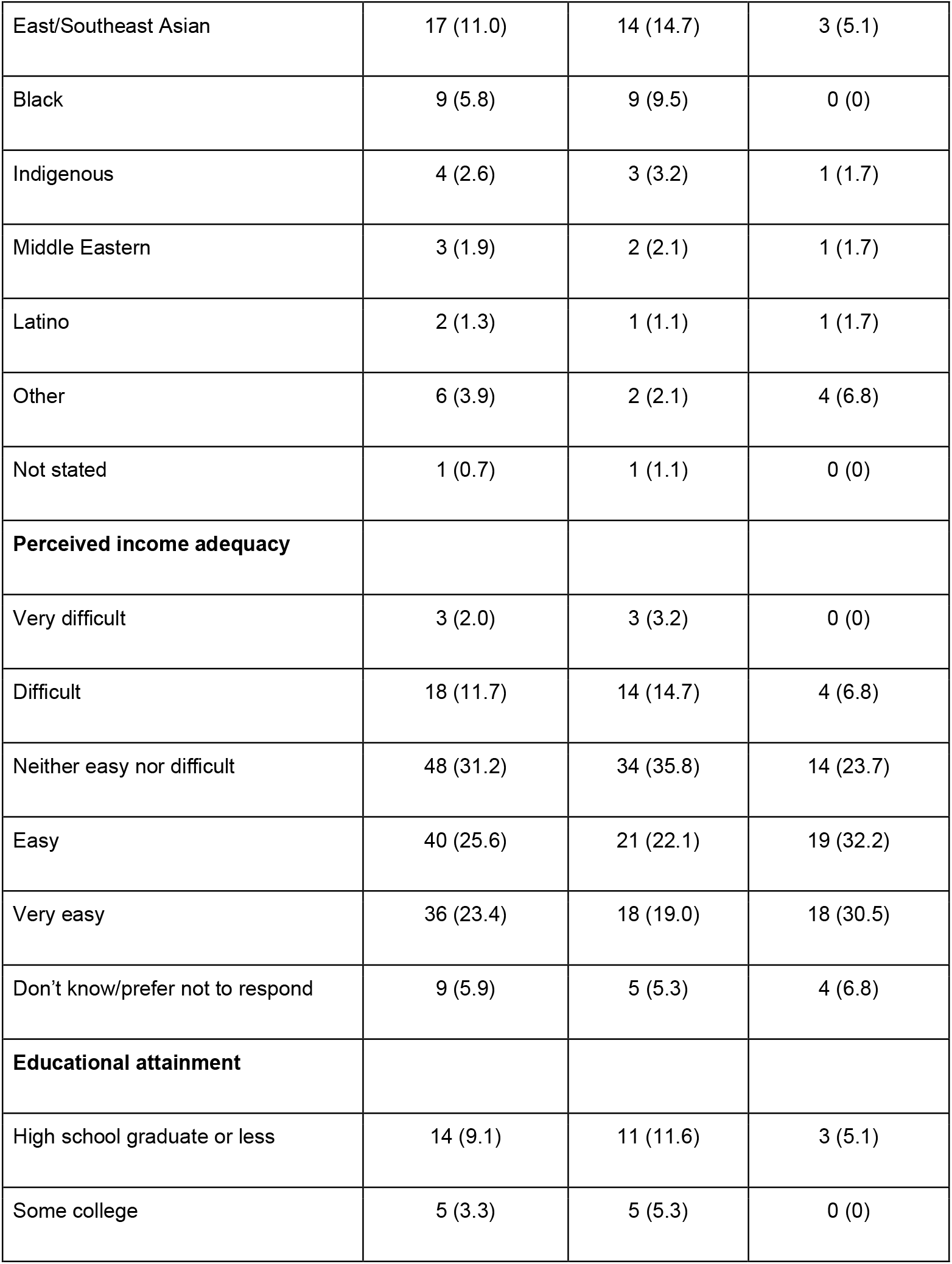
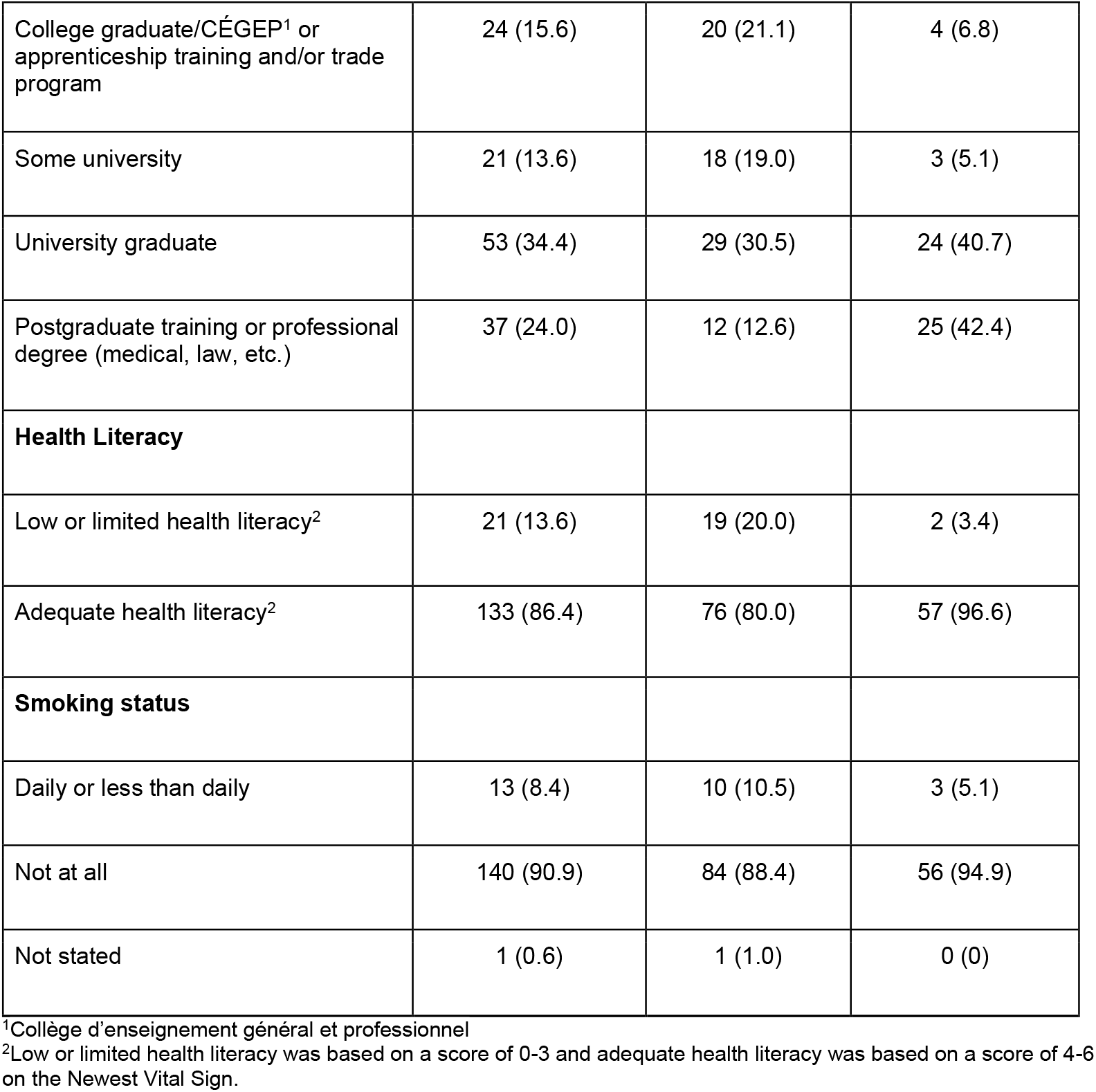
Sociodemographic and health characteristics of adults (n=154) participating in a study to evaluate the construct validity of the Canadian Food Intake Screener.

The mean screener score was 35.0 (SD=4.7) of 65 possible points, ranging from 25.1 at the 1^st^ percentile to 45.2 at the 99^th^ percentile (**Figure 2**). There was moderate to strong evidence of differences in screener scores among subgroups in hypothesized directions (**Table 4**) by gender identity (p=0.06), perceived income adequacy (p=0.07), educational attainment (p=0.02), and smoking status (p=0.003). Differences were not observed by age group or health literacy.

**Table 4:** Comparison of mean Canadian Food Intake Screener scores, of a possible 65 points, among subgroups known to differ in dietary quality among adults 18-65 years (n=154)

| Characteristic | Mean Screener Score | Standard Deviation | P-value<sup>1</sup> |
| --- | --- | --- | --- |
| **Age** |  |  |  |
| 18-24 (n=26) | 34.8 | 4.8 | 0.88 |
| 25-34 (n=45) | 35.1 |  |  |
| 35-44 (n=46) | 34.5 |  |  |
| 45-54 (n=22) | 35.3 |  |  |
| 55-65 (n=15) | 36.2 |  |  |
| **Gender<sup>2</sup>** |  |  |  |
| Men (n=38) | 33.8 | 4.7 | 0.06 |
| Women (n=114) | 35.4 |  |  |
| **Perceived income adequacy<sup>3</sup>** |  |  |  |
| Difficult or very difficult (n=21) | 33.0 | 4.7 | 0.07 |
| Neither easy nor difficult (n=48) | 34.7 |  |  |
| Easy or very easy (n=76) | 35.5 |  |  |
| **Educational attainment** |  |  |  |
| College graduate/CÉGEP<sup>4</sup> or apprenticeship training and/or trade program (n=43) | 33.6 | 4.7 | 0.02 |
| Some university or greater (n=111) | 35.6 |  |  |
| **Health literacy<sup>5</sup>** |  |  |  |
| Low or limited health literacy (0-3) (n=21) | 33.9 | 4.7 | 0.22 |
| Adequate health literacy (4-6) (n=133) | 35.2 |  |  |
| **Smoking status<sup>6</sup>** |  |  |  |
| Daily or less than daily (n=13) | 30.5 | 4.6 | 0.003 |
| Not at all (n=140) | 35.4 |  |  |

<sup>1</sup>P-values were derived using one-way ANOVA and applying Levene's test for homogeneity to determine whether a Welch's ANOVA was appropriate in the cases in which the assumption of homogeneity of variances was not met.
<sup>2</sup>Two individuals were not included because they identified as another gender identity or did not provide details on their gender identity.
<sup>3</sup>Nine individuals were not included because they had missing data on perceived income adequacy.
<sup>4</sup>Collège d'enseignement général et professionnel
<sup>5</sup>Low or limited health literacy was defined as a score of 0-3 and adequate health literacy was defined as a score of 4-6 on the Newest Vital Sign tool (Weiss et al. 2005).
<sup>6</sup>One individual was not included because they did not state their smoking status.

**Figure 2:**
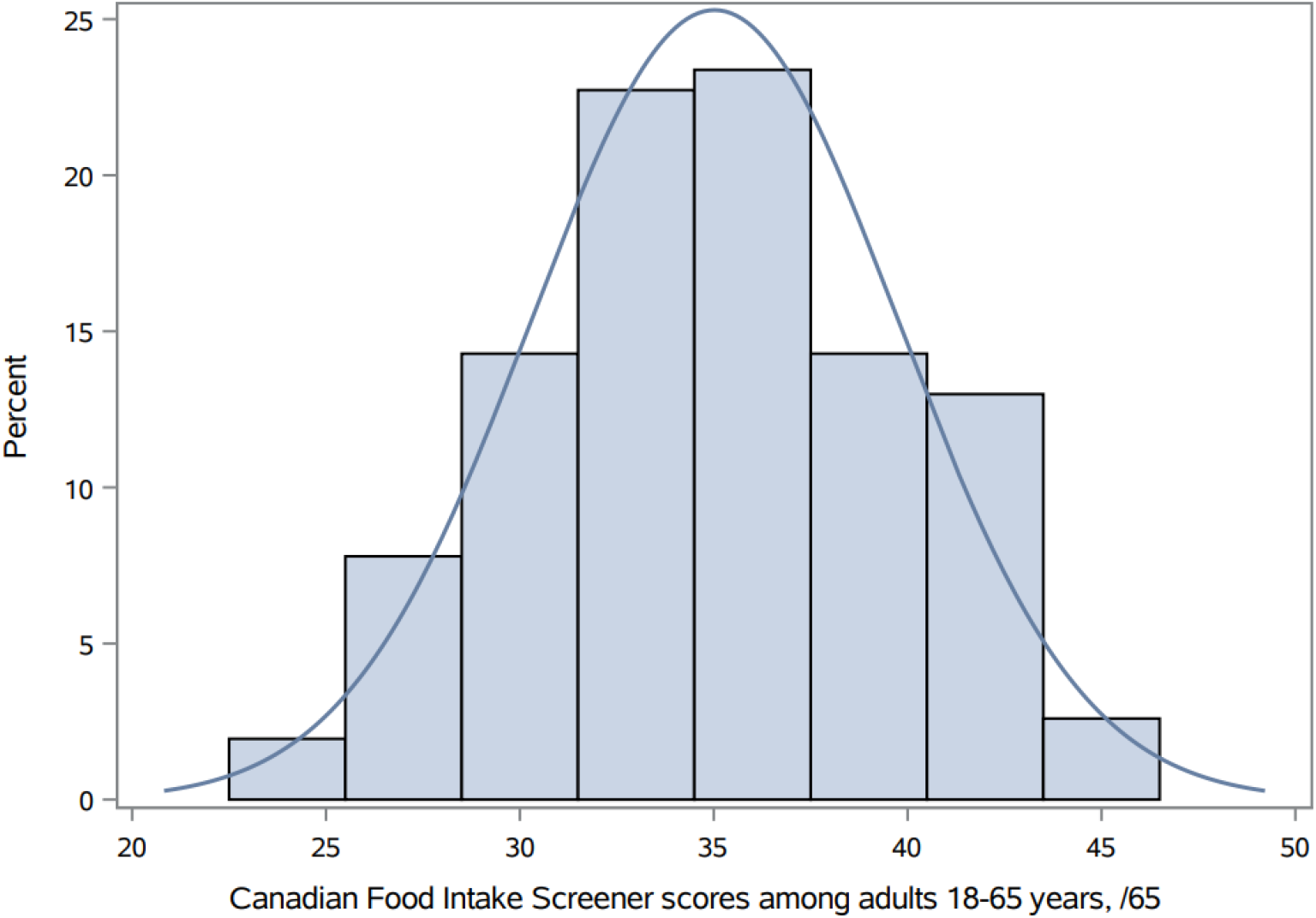
Distribution of Canadian Food Intake Screener scores among adults aged 18-65 years (n=154) in a study to examine the construct validity of the screener.

The mean HEFI-2019 score based on simulated usual intakes was 40.9 (SE=1.2) of 80 possible points, ranging from 25.9 at the 1^st^ percentile to 56.0 at the 99^th^ percentile (**Figure 3**). A moderately strong Pearson correlation coefficient of 0.53 (SE=0.12) was observed between screener scores and total HEFI-2019 scores. Total screener scores explained 28% (SE=12%) of the variability in total HEFI-2019 scores. The correlation between screener scores and simulated usual energy intake was 0.08 (SE=0.09).When the scores for the screener components generated to arrive at the total screener score were included in the model in place of the total score, they collectively explained 53% (SE=9.9%) of the variability in total HEFI-2019 scores.

**Figure 3:**
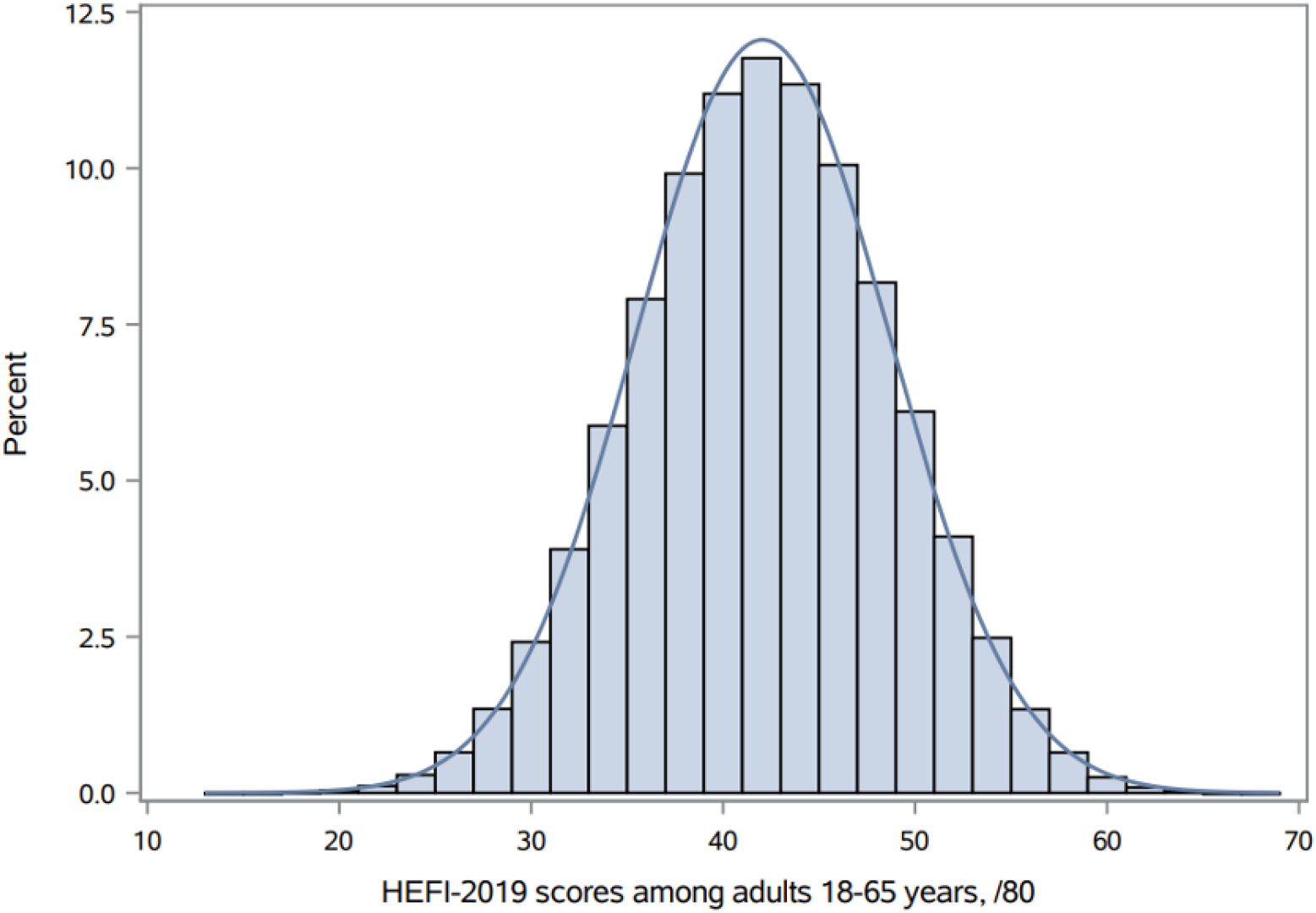
Distribution of total Healthy Eating Food Index-2019 scores^1^ among adults aged 18-65 years in a study to examine the construct validity of the Canadian Food Intake Screener. ^1^ Healthy Eating Food Index-2019 scores were based on simulated usual intakes, using data from up to two non-consecutive 24-hour dietary recalls (n=230 recalls completed by 128 individuals).

When truncation was applied in generating screener scores, the correlation coefficient and proportion of variability in total HEFI-2019 scores explained were slightly lower (data not shown). Similarly, when a scoring approach that did not incorporate summing across questions or deriving ratios to create components was used, the correlation coefficient and proportion of variability explained were lower compared to those based on the application of the scoring system (data not shown).

## Discussion

The Canadian Food Intake Screener was developed to rapidly assess the overall alignment of adults’ dietary intake with the CFG-2019 healthy food choices recommendations. The present findings suggest moderate construct validity of the screener, supporting its use to provide rapid insights into overall alignment of intake over the past month with the CFG-2019 healthy food choices recommendations when comprehensive dietary assessment is not possible. The mean screener score in the sample was 35.0 of 65 possible points and reasonable variation was observed. The 99^th^ percentile was 45.2, indicating considerable room for improvement with respect to alignment of intakes with the recommendations. Similarly, the 99^th^ percentile for HEFI-2019 scores was 63 of 80 possible points based on analysis of the 2015 CCHS-Nutrition data (Brassard et al. 2022b) and 56 in the current sample.

The correlation coefficient of 0.53 between screener and total HEFI-2019 scores is similar to that observed for other brief screeners, relative to data from more comprehensive dietary assessment instruments. For example, correlation coefficients between Behavioral Risk Factor Surveillance System fruit and vegetable module data and intake data from food frequency questionnaires, 24HDRs, and food records range from 0.29 to 0.63 (Centers for Disease Control and Prevention n.d.). An evaluation of a 40-item screener to assess alignment with the Dutch food-based dietary guidelines found a Kendall’s tau-b coefficient of 0.51 in a sample of 751 adults, based on comparing screener scores to Dutch Healthy Diet index 2015 scores derived from a food frequency questionnaire (de Rijk et al. 2021). Similarly, an evaluation of the 14-item Mediterranean Diet Adherence Screener among a sample of 7146 adults found a Pearson’s correlation coefficient of 0.52 with scores indicating adherence to the Mediterranean diet based on a food frequency questionnaire (Schröder et al. 2011). Among a sample of 50 college students, a Pearson’s correlation coefficient of 0.79 was observed between scores on the 22-item Short Healthy Eating Index and Healthy Eating Index-2015 scores, estimated using repeat 24HDRs (Colby et al. 2020). This relatively high correlation for the latter screener may relate, in part, to the more homogenous population of students and the underlying quantitative recommendations, including recommended amounts to consume daily.

The mean total HEFI-2019 score in the sample was 40.9, compared to 43.0 based on the 2015 CCHS-Nutrition, which included individuals aged two and up (Brassard et al. 2022b), lending confidence to the present study and its findings. The distribution of total HEFI-2019 scores in this sample was somewhat narrower, possibly due to the restricted age range, as well as the lack of national representation and less heterogeneity. The largest difference in screener scores observed between subgroups was five points, between those who did and did not report smoking. Similarly, in the evaluation of the HEFI-2019 using data from the 2015 CCHS-Nutrition, the corresponding difference in total HEFI-2019 scores was seven points (Brassard et al., 2022b). Differences were not observed in screener scores by age, possibly due to the categorical nature of the age variable and the small cell sizes for some categories. The sample size and skew toward higher perceived income adequacy and higher health literacy may also underlie the lack of meaningful differences by these characteristics.

The HEFI-2019 (Brassard et al. 2022a, 2022b) and the Healthy Eating Index-2015 (Krebs-Smith et al. 2018) are density-based, helping to decouple diet quality from quantity. In contrast, the focus on frequency of consumption within the screener and its scoring system raises the possibility that those with higher energy requirements and intakes, particularly of healthy foods, may receive higher screener scores. Within this sample, there was no meaningful correlation between screener scores and energy intake based on the 24HDRs. However, the measurement error in estimated energy intake based on self-report methods (Freedman et al. 2014; Kirkpatrick et al. 2022b) makes it challenging to detect an association between screener scores and true usual energy intake. Because the CFG-2019 does not provide quantitative recommendations for food categories, no upper limit on frequency was applied in determining the standards for maximum scores for healthy foods and minimum scores for foods to limit.

Truncation was considered to relax the standards but did not improve the correlation coefficient with total HEFI-2019 scores, perhaps because it introduces its own problems, such as floor or ceiling effects (Ricart et al. 2022). Alternative assessment approaches, such as images of plates captured using mobile food records (Boushey et al. 2017; Ho et al. 2020) and analyzed using machine learning, may allow for more direct consideration of relative intake of different food categories in the future, consistent with the focus of the CFG-2019 plate on proportionality (Health Canada, 2022). However, while potentially reducing the burden for respondents, like other methods of dietary assessment, image-based technology is subject to measurement error (Ho et al. 2020).

While dietary screeners are not a replacement for comprehensive dietary assessment, their simplicity makes them a popular choice among researchers and practitioners (Kirkpatrick et al. 2014a, 2017; Rodrigo et al. 2015). The Canadian Food Intake Screener can be self-administered in approximately five minutes and was designed to be as simple as possible, though it requires recalling and averaging frequency of intake over the past month, which can be cognitively challenging (National Cancer Institute 2015). In contrast, 24HDRs integrate multiple passes designed to prompt respondents to report their intake for the prior day in as much detail as possible. Although web-based self-administered systems, such as ASA24 (Subar et al. 2012) and Rappel 24h Web (R24W) (Jacques et al. 2016; Lafrenière et al. 2017), make it possible to collect 24HDRs in a range of contexts, a recall takes 20 to 30 minutes or more to complete, and non-consecutive repeat administrations are needed on at least a subsample to enable the estimation of distributions of usual intake (Dodd et al. 2006). Further, cleaning the recalls and preparing the variables to calculate HEFI-2019 scores in the current study required >75 hours of a registered dietitian’s time. These steps are less burdensome when using secondary data that have been cleaned and can be readily linked with information on reference amounts and free sugars. Nonetheless, the screener scoring system is, by design, simpler than the HEFI-2019 scoring algorithm.

Decisions about whether to use the screener versus the HEFI-2019 should be informed not only by considerations of resources and burden, but more importantly, by the research question. Scores based on the screener do not consider all components assessed by the HEFI-2019 (Brassard et al. 2022a, 2022b). Because the screener does not assess water consumption (Hutchinson et al. 2022), a beverages component is not included in the screener scoring system. Furthermore, the screener assesses frequency of consumption of key sources of added sugars (Mela and Woolner, 2018), saturated fat, and sodium, whereas HEFI-2019 scoring involves quantification of intake of free sugars, saturated and unsaturated fat, and sodium (Brassard et al. 2022b). Further, component scores used to calculate total screener scores should not be interpreted individually because they are based on one or a few questions that do not provide sufficient information to quantify total intake of a specific group of foods, such as fruits and vegetables. Rather, the total screener score is intended to signal higher or lower degrees of overall alignment with the CFG-2019 healthy food choices recommendations. In contrast, given the underlying detailed intake data, the HEFI-2019 enables consideration of individual component scores, with interpretation of component scores recommended to contextualize total HEFI-2019 scores (Brassard et al. 2022a, 2022b).

The screener could be administered in national surveillance to provide high-level insights on the alignment of adults’ intake with the CFG-2019 healthy food choices recommendations.

However, it should not replace the collection of 24HDR data, including in periodic nutrition-focused cycles of the CCHS (Health Canada 2004, 2015). Compared to the screener, 24HDR data are more accurate and allow for richer analyses (Dodd et al. 2006). In addition to the calculation of HEFI-2019 total and component scores (Brassard et al. 2022a, 2022b), such analyses may estimate the prevalence of inadequate or excess nutrient intakes relative to the Dietary Reference Intakes (Institute of Medicine 2006), identify top sources of food categories and nutrients of public health concern, and characterize meal patterns and associated contextual factors.

The use of the screener to evaluate interventions requires careful interpretation since exposure to an intervention can change reporting of dietary intake (Neuhouser et al. 2008; Natarajan et al. 2010), though this may be less of a concern for interventions focused on environmental factors versus individual practices (National Cancer Institute 2015). As well, the level of change that the screener can detect has not been assessed. Relatedly, consideration of what constitutes meaningful differences in screener scores over time or among groups is needed. For the HEI, which is scored to a maximum of 100 points to assess alignment of intakes with the U.S. Dietary Guidelines for Americans, it has been suggested that differences of 5-6 points between independent groups might be meaningful (Kirkpatrick et al. 2018). There is no threshold at which screener scores are indicative of meeting the CFG-2019 healthy food choices recommendations or of a ‘healthy’ diet. For knowledge translation purposes, scores can be categorized to describe the degree of alignment with the guidance (e.g., higher or lower than the mean or median in a given sample). However, scores should not be categorized for analysis purposes as this results in loss of information and potentially, misclassification of values near the thresholds (Kirkpatrick et al. 2018).

In the current sample, the correlation between scores based on the screener scoring system and total HEFI-2019 scores was somewhat higher compared to that for an approach that did not involve the creation of components and weighting, indicating that the scoring system adds value in aligning screener scores with the CFG-2019 healthy food choices recommendations. However, when scores assigned to separate screener components were considered in place of total screener scores, a larger proportion of variability in total HEFI-2019 scores was explained. Though the addition of independent variables to a model results in a higher R-squared estimate by definition, this finding suggests that the scoring algorithm could potentially be further optimized. Administration of the screener, potentially along with 24HDRs, in larger samples may enable refinement of the scoring system using data-driven approaches.

For example, in the development of the scoring approach for the Short Healthy Eating Index, Colby et al. (2020) drew upon data from 398 college students to identify the best predictors of diet quality using classification and regression tree algorithms. Lafrenière et al. (2019) also used a classification and regression tree approach to develop a brief measure of diet quality, drawing upon pooled data from approximately 5000 adults who completed a food frequency questionnaire as part of several studies. Drawing upon 24HDR data for approximately 7600 individuals participating in the 2009-2010 National Health and Nutrition Examination Survey (86% of whom completed two recalls), Thompson et al. (2017) developed scoring algorithms to align estimates from the Dietary Screener Questionnaire more closely with those from 24HDRs. However, Gilsing et al. (2018) explored the development of scoring algorithms for the screener used in the Canadian Longitudinal Study on Aging, using data from 200 older adults who completed four 24HDRs, and found the algorithms did not notably improve mean nutrient intake estimates. Additionally, data-driven approaches based on one dataset may not be portable to other datasets, for example, due to differences in sociodemographic characteristics or the time periods during which data were collected (Thompson et al. 2017). Further, given their limitations, there is a ceiling on the extent to which screeners and their scoring systems can be improved.

To improve the consistency of periods captured by the screener and the HEFI-2019 scores, modeling was used to estimate total HEFI-2019 scores based on the simulated distribution of usual intakes (Dodd et al. 2006, Brassard et al. 2022b). A similar approach has been used in other validation research (Colby et al. 2020, Gilsing et al. 2018; Hewawitharana et al. 2018; Thompson et al. 2017). Other methods, such as Bland-Altman analysis (Giavarina, 2015) to assess agreement between estimates from two error-prone instruments (Alcantara et al. 2015; de Rijk et al. 2021; Schröder et al. 2011, 2012), were not appropriate because the modeling used to produce HEFI-2019 scores does not produce an individual-level analogue to the screener score (Kirkpatrick et al. 2018, 2022a). While the screener was evaluated among adults aged 18-65 years, it could be evaluated and/or adapted for use with other populations, such as youth and older adults, as described in the accompanying paper (Hutchinson et al. 2022).

Several additional considerations are relevant to the interpretation of our findings. The collection of two non-consecutive 24HDRs enabled comparison of screener scores to total HEFI-2019 scores based on the simulated distribution of usual intakes, which represented the strongest available reference measure for assessment of the screener’s construct validity since no objective marker of usual diet quality is known. Multivariate usual intake modeling accounted for within-person random error, as well as the multiple correlated dietary components considered in scoring the HEFI-2019 (Brassard et al. 2022b). However, 24HDR data are affected by bias (Freedman et al. 2014, 2015; Kirkpatrick et al. 2022b), and the correlation between screener and HEFI-2019 scores may be overestimated due to correlated errors between the screener and 24HDR data (Kirkpatrick et al. 2019a). The sample of 154 was close to the desired sample size; and quotas for language, race/ethnicity, and educational attainment were achieved. However, the sample was skewed towards women (with 25% identifying as men, compared to the target of 30%), and 24HDRs were available for a subset of participants. The smaller than intended sample with recall data limits the precision of the observed correlation between screener and HEFI-2019 scores. The overall sample size also did not support stratification of analyses by language and other characteristics.

Bogus participants are adaptive to quotas and other attempts at dissuasion (Almazyad et al. 2011; Shamon and Berning 2019; Pei et al. 2020; Wang et al. 2021). Intensive data quality appraisal and cleaning were implemented to ensure data from bogus participants were not included in the analyses. Data collection was conducted from July to December, though each participant completed their study tasks within a one-month period, and few effects of seasonality on dietary intake are apparent within the North American context (Bernstein et al. 2016). Finally, testing of the screener was conducted online, and effects related to mode of administration are possible (Bowling 2005).

## Conclusions

The results of this study suggest moderate construct validity of the Canadian Food Intake Screener, supporting its use to rapidly assess overall alignment of adults’ intake over the past month with the CFG-2019 healthy food choices recommendations. Use of the screener in research and surveillance, as appropriate, can contribute to a robust evidence base that can be synthesized to inform policies and programs to improve dietary quality. Administration of the screener to large, diverse samples in the future may enable refinement of the screener’s scoring system to improve its alignment with the underlying recommendations.

## Data Availability

Data are available from the authors upon request.

## Acknowledgements

The authors are grateful to the participants in the construct validity study, and to Kimberley J. Hernandez, Isabelle Rondeau, Lisa-Anne Elvidge Munene, Sylvie St-Pierre, and other Health Canada employees who participated in and provided input on various aspects of the project. The authors are also grateful to Dr. Didier Brassard from McGill University (formerly of Université Laval) for providing advice on the application of HEFI-2019 and Sanaa Hussain from the University of Waterloo for support with data cleaning and creation of the code book, as well as referencing.

## Contributions

Conception and design: SIK, JMH, KWD, PMG, BL, JH, AW; analysis and interpretation of data: JMH, SIK, KWD, PMG, BL; drafting of the manuscript: JMH, SIK; revising the manuscript critically for important intellectual content: KWD, PMG, BL, JH, AW, MP, TEW, MLCL, MJ, SL, DLO, RP, JRS, JEV, KS, IR; final approval of the manuscript submitted: all authors.

## Data availability

Data are available from the authors upon request.

