## Supplemental File S2 for "The Canadian Food Intake Screener for assessing alignment of adults’ dietary intake with the 2019 Canada’s Food Guide healthy food choices recommendations: Scoring system and construct validity"

Hutchinson JM, et al.

**Supplementary File S2. Expert advisors who provided input on the development of the Canadian Food Choices Screener**

- Meghan Day, British Columbia Ministry of Health
- Kevin Dodd, U.S. National Cancer Institute
- Patricia Guenther, University of Utah
- Jess Haines, University of Guelph
- Mahsa Jessri, University of British Columbia
- Mary L'Abbé, University of Toronto
- Benoît Lamarche, Université Laval
- Simone Lemieux, Université Laval
- Maria Laura Louzada, University of São Paulo
- Dana Lee Olstad, University of Calgary
- Rachel Prowse, Memorial University
- Janis Randall Simpson, University of Guelph
- Jill Reedy, U.S. National Cancer Institute
- Hassan Vatanparast, University of Saskatchewan
- Jennifer Vena, Alberta Health Services
