## Supplemental File S3 for "The Canadian Food Intake Screener for assessing alignment of adults’ dietary intake with the 2019 Canada’s Food Guide healthy food choices recommendations: Scoring system and construct validity"

Hutchinson JM, et al.

**Supplementary File S3. Canadian Food Intake Screener/Questionnaire court canadien sur les apports alimentaires: Scoring System READ ME File**

### READ ME

#### Background

The Canadian Food Intake Screener/Questionnaire court canadien sur les apports alimentaires was developed to rapidly assess alignment of adults' dietary intake over the past month with the 2019 Canada Food Guide's healthy food choices recommendations. The screener includes 16 questions. Arriving at screener scores involves creating components by summing and deriving ratios based on responses to screener questions. Scores on each component are summed to arrive at a total possible score with a maximum of **65 points**.

The screener is designed to provide one total score assessing alignment. The component scores are meant only for the purpose of arriving at total scores.

Details on the scoring system can be found in the accompanying manuscript (Hutchinson et al. 2022).

#### Suggested layout for input dataset

The sample SAS code was created with the assumption that observations are rows and screener responses are columns.

#### Sample code

Sample SAS code is provided in the accompanying supplementary file (Supplementary File S4), demonstrating how components used in arriving at screener scores are created and summed.

#### Output

From the sample SAS code, several variables are created:

- Intermediate variables used to create components for scoring:
    - CFIS_FVSUM
    - CFIS_PROT_TOT
    - CFIS_PB_RATIO
    - CFIS_GR_TOT
    - CFIS_GR_RATIO
    - CFIS_4S
    - CFIS_5S
    - CFIS_10S
    - CFIS_11S

    - CFIS_12S
    - CFIS_13S
- Component variables:
    - CFIS_FV = vegetables and fruit component
    - CFIS_PB_PRO = plant-based protein: total protein component
    - CFIS_PROT_SUM = total protein component
    - CFIS_WG = whole grain component
    - CFIS_GR_Score = whole grain: total grain component
    - CFIS_SW_Score = sugary foods component
    - CFIS_SALT_Score = salty foods component
    - CFIS_OIL = unsaturated oils component
- Total score:
    - CFIS_Total = total screener score
