## Supplemental File S4 for "The Canadian Food Intake Screener for assessing alignment of adults’ dietary intake with the 2019 Canada’s Food Guide healthy food choices recommendations: Scoring system and construct validity"

```
**************************************************************************************************************;
**************************************************************************************************************;
***The Canadian Food Intake Screener for assessing alignment of adults’ dietary intake with the 2019********************;
***Canada’s Food Guide healthy food choices recommendations: Scoring system and construct validity**********************;

***Hutchinson JM, et al.*************************************************************************************;

***Supplementary File S4. Sample Code to Score the Canadian Food Intake Screener****************************************;
**************************************************************************************************************;
**************************************************************************************************************;

*NOTE: Components are intended only for the purpose of summing to achieve a total score;

*Screener variable names:;
*CFIS_1:  Fruits;
*CFIS_2:  Potatoes;
*CFIS_3:  Vegetables;
*CFIS_4:  Fast food;
*CFIS_5:  Processed meat;
*CFIS_6:  Animal-based protein foods;
*CFIS_7:  Plant-based protein foods;
*CFIS_8:  Yogurt, kefir, cheese;
*CFIS_9:  Unsweetened milk;
*CFIS_10: Sweetened milk;
*CFIS_11: Sweetened beverages;
*CFIS_12: Sugary snacks;
*CFIS_13: Salty snacks;
*CFIS_14: Refined grain foods;
*CFIS_15: Whole grain foods;
*CFIS_16: Unsaturated oils;
```

```
data samplescoring;
set inputdata;

***Vegetables and fruit***;
*Sum vegetable + potato + fruit items
*Weight to /20;
CFIS_FVSUM=(CFIS_1+CFIS_2+CFIS_3);
CFIS_FV=CFIS_FVSUM/1.35;

***Proteins***;
*Score comprised of two components: the ratio of;
*plant-based protein foods/total protein and total protein;


*Ratio of plant-based protein foods item/total proteins (sum of animal-based protein foods,
*plant-based protein foods, yogurt, kefir, cheese, and unsweetened milk);
*Weight to /5;
CFIS_PROT_TOT = (CFIS_6 + CFIS_7 + CFIS_8 + CFIS_9);
CFIS_PB_RATIO = CFIS_7/CFIS_PROT_TOT;
if CFIS_PB_RATIO =1 then CFIS_PB_PRO = 5;
else if CFIS_PB_RATIO = 0 then CFIS_PB_PRO = 0;
else if CFIS_PB_RATIO >0 & CFIS_PB_RATIO <1 then CFIS_PB_PRO=5*(CFIS_PB_RATIO);

*Total protein (sum of animal-based protein foods, plant-based protein foods,;
*yogurt, kefir, cheese, and unsweetened milk);
*Weight to /5;
CFIS_PROT_SUM = CFIS_PROT_TOT/7.2;

***Whole grains***;
*Score comprised of two components: total whole grain and the ratio;
*of whole-grain foods over total grains (whole-grain foods plus refined-grain foods);

*Total whole-grain foods;
*Weight to /5;
CFIS_WG = CFIS_15/1.8;

*Grain foods ratio (Whole-grain foods/(Whole-grain foods + Refined-grain foods));
*Weight to /5;
CFIS_GR_TOT = CFIS_14 + CFIS_15;
if CFIS_GR_TOT ne 0 then CFIS_GR_RATIO = CFIS_15/CFIS_GR_TOT;
    else if CFIS_GR_TOT = 0 then CFIS_GR_RATIO = 0;
if CFIS_GR_RATIO = 0 then CFIS_GR_Score = 0;
    else if CFIS_GR_RATIO =1 then CFIS_GR_Score = 5;
    else if CFIS_GR_RATIO >0 & CFIS_GR_RATIO <1 then CFIS_GR_Score=5*(CFIS_GR_RATIO);
```

```
***Foods and beverages high in sugars***;
*Weight to /10;
*Foods to limit are reverse-scored;

*scoring sweetened milk (reverse), CFIS_10;
    if CFIS_10 = 0 then CFIS_10S = 9;
    else if CFIS_10 = 1 then CFIS_10S = 8;
    else if CFIS_10 = 2 then CFIS_10S = 7;
    else if CFIS_10 = 3 then CFIS_10S = 6;
    else if CFIS_10 = 4 then CFIS_10S = 5;
    else if CFIS_10 = 5 then CFIS_10S = 4;
    else if CFIS_10 = 6 then CFIS_10S = 3;
    else if CFIS_10 = 7 then CFIS_10S = 2;
    else if CFIS_10 = 8 then CFIS_10S = 1;
    else if CFIS_10 = 9 then CFIS_10S = 0;
    else CFIS_10S=".";
*scoring sweetened beverages (reverse), CFIS_11;
    if CFIS_11 = 0 then CFIS_11S = 9;
    else if CFIS_11 = 1 then CFIS_11S = 8;
    else if CFIS_11 = 2 then CFIS_11S = 7;
    else if CFIS_11 = 3 then CFIS_11S = 6;
    else if CFIS_11 = 4 then CFIS_11S = 5;
    else if CFIS_11 = 5 then CFIS_11S = 4;
    else if CFIS_11 = 6 then CFIS_11S = 3;
    else if CFIS_11 = 7 then CFIS_11S = 2;
    else if CFIS_11 = 8 then CFIS_11S = 1;
    else if CFIS_11 = 9 then CFIS_11S = 0;
    else CFIS_11S=".";
*scoring sugary treats (reverse), CFIS_12;
    if CFIS_12 = 0 then CFIS_12S = 9;
    else if CFIS_12 = 1 then CFIS_12S = 8;
    else if CFIS_12 = 2 then CFIS_12S = 7;
    else if CFIS_12 = 3 then CFIS_12S = 6;
    else if CFIS_12 = 4 then CFIS_12S = 5;
    else if CFIS_12 = 5 then CFIS_12S = 4;
    else if CFIS_12 = 6 then CFIS_12S = 3;
    else if CFIS_12 = 7 then CFIS_12S = 2;
    else if CFIS_12 = 8 then CFIS_12S = 1;
    else if CFIS_12 = 9 then CFIS_12S = 0;
    else CFIS_12S=".";


CFIS_SW_SCORE=(CFIS_10S + CFIS_11S + CFIS_12S)/2.7;

***Foods high in sodium/saturated fat***;
*Weight to /10;
*Foods to limit are reverse-scored;

*scoring fast food (reverse), CFIS_4;
    if CFIS_4 = 0 then CFIS_4S = 9;
    else if CFIS_4 = 1 then CFIS_4S = 8;
    else if CFIS_4 = 2 then CFIS_4S = 7;
    else if CFIS_4 = 3 then CFIS_4S = 6;
    else if CFIS_4 = 4 then CFIS_4S = 5;
    else if CFIS_4 = 5 then CFIS_4S = 4;
    else if CFIS_4 = 6 then CFIS_4S = 3;
    else if CFIS_4 = 7 then CFIS_4S = 2;
    else if CFIS_4 = 8 then CFIS_4S = 1;
    else if CFIS_4 = 9 then CFIS_4S = 0;
    else CFIS_4S=".";
*scoring processed meat (reverse), CFIS_5;
    if CFIS_5 = 0 then CFIS_5S = 9;
    else if CFIS_5 = 1 then CFIS_5S = 8;
    else if CFIS_5 = 2 then CFIS_5S = 7;
    else if CFIS_5 = 3 then CFIS_5S = 6;
    else if CFIS_5 = 4 then CFIS_5S = 5;
    else if CFIS_5 = 5 then CFIS_5S = 4;
    else if CFIS_5 = 6 then CFIS_5S = 3;
    else if CFIS_5 = 7 then CFIS_5S = 2;
    else if CFIS_5 = 8 then CFIS_5S = 1;
    else if CFIS_5 = 9 then CFIS_5S = 0;
    else CFIS_5S=".";
*scoring salty snacks (reverse), CFIS_13;
    if CFIS_13 = 0 then CFIS_13S = 9;
    else if CFIS_13 = 1 then CFIS_13S = 8;
```

```
    else if CFIS_13 = 2 then CFIS_13S = 7;
    else if CFIS_13 = 3 then CFIS_13S = 6;
    else if CFIS_13 = 4 then CFIS_13S = 5;
    else if CFIS_13 = 5 then CFIS_13S = 4;
    else if CFIS_13 = 6 then CFIS_13S = 3;
    else if CFIS_13 = 7 then CFIS_13S = 2;
    else if CFIS_13 = 8 then CFIS_13S = 1;
    else if CFIS_13 = 9 then CFIS_13S = 0;
    else CFIS_13S=".";

CFIS_SALT_SCORE=(CFIS_4S + CFIS_5S + CFIS_13S)/2.7;

***Unsaturated oils***;
*Weight to /5;

CFIS_OIL = CFIS_16/1.8;

*delete unnecessary variables;
delete CFIS_FVSUM CFIS_PROT_TOT CFIS_PB_RATIO CFIS_GR_TOT CFIS_GR_RATIO;

***Sum components to get total score***;
CFIS_TOTAL= (CFIS_FV + CFIS_PB_PRO + CFIS_PROT_SUM + CFIS_WG + CFIS_GR_Score +  CFIS_SW_SCORE + CFIS_SALT_SCORE + CFIS_OIL);

run;
```
